# Default Mode Network Detection using EEG in Real-time

**DOI:** 10.1101/2024.04.03.24305235

**Authors:** Navin Cooray, Chetan Gohil, Brendan Harris, Shaun Frost, Cameron Higgins

## Abstract

Mental health disorders affect countless people worldwide and present a major challenge for mental health services, which are struggling with the demand on a global scale. Recent studies have indicated that activity of the brain’s Default Mode Network (DMN) could prove insightful in monitoring patient recovery from depression and has been used as a therapeutic target itself. An opportunity exists to replicate recent therapeutic protocols targeting DMN connectivity via functional magnetic resonance imaging using the more economically scalable modality of electroencephalogram (EEG). The aim of this work was to validate the accuracy of real-time DMN detection methods applied to EEG data, using a publicly available dataset. Using a Hidden Markov Model to identify a 12-state resting-state network, this work achieved an overall DMN detection accuracy of 95%. Furthermore, the model was able to achieve a correlation of 0.617 between the baseline and calculated DMN fractional occupancy. These results demonstrate the ability of real-time analysis to effectively identify the DMN through EEG data providing an avenue for further applications that monitor and treat mental health disorders.

## I. Introduction

The DMN describes a set of brain regions that synchronously coactivate during ‘idle’ periods of wakefulness. This brain network has functional roles supporting attention and cognition [1], [2], and is associated with a number of introspective cognitive states, including episodic memory and future-oriented thought [3], [4]. The clinical significance and importance of the DMN continues to grow in scientific literature, with DMN dysfunction associated with numerous mental health disorders, including depression [5], [6]. Normalisation of DMN connectivity is theorised to be the mechanism of action of transcranial magnetic stimulation (TMS), a brain stimulation therapy that has proven effective in treating depression [6]. Neurofeedback methods implemented using functional magnetic resonance (fMRI) imaging to target DMN connectivity patterns have also been shown to reduce depressive symptoms in patients [7], [8].

Methods that detect DMN activity have been widely implemented in fMRI, but have also been demonstrated in non-invasive neurophysiology modalities such as EEG [9]. Whereas DMN-neurofeedback methods implemented in fMRI have been effective in reducing depressive symptoms [7],[8], these methods face significant economic barriers to becoming a scalable therapy. EEG based DMN-Neurofeedback has the potential to achieve comparable results whilst presenting drastically fewer practical and economic barriers to patient adoption. This could facilitate a quantitative and data-driven approach to mental healthcare, by augmenting current therapeutic practices with real-time readouts of DMN activity changes that assess and assist in clinical decisions and interventions. There is a clear motivation and increasing need to quantify, define, and understand the DMN.

In this work, we compare a commercially available software package for real-time DMN detection against an emerging gold-standard methodology used in the research space for post-hoc inference of DMN activity. Applied in an offline fashion and utilising a number of techniques for noise reduction and source space signal estimation that are not technically feasible in real-time, this emerging gold-standard has been used to characterise physiological [10], cognitive [11] and task-based [12] characteristics of DMN activity. If validated, the ability to monitor this network in real-time at the high temporal resolution of EEG would present new opportunities for clinical monitoring and intervention in mental health illnesses.

## II. Data

The Leipzig Study for Mind-Body-Emotions Interactions (LEMON) dataset consists of 227 participants with two distinct age groups, a young cohort (N=153, 25.1±3.1 years, 45 female) and an elderly cohort (N=74, 67.6±4.7 years, 37 female), sourced from Leipzig, Germany between 2013 and 2015 [13]. The study protocol and open access publication of the data for research purposes was approved by the ethics committee at the medical faculty of the University of Leipzig (reference number 154/13-ff), with all participants providing consent to their data being shared anonymously prior to any data acquisition. The dataset includes a resting-state fMRI and a 62-channel EEG experiment taken at rest (N=216) over 16 min. The EEG recording consists of 16 blocks, each 60s long, eight with eyes-closed (EC) and eight with eyes-open (EO), where EC and EO were interleaved. Participants were asked to stay awake while fixating eyes on a computer screen. For the purposes of this study, the subset of 110 subjects was selected for whom both fMRI structural scans and EEG channel localizer data was available.

## III. Methodolodgy

In this work we measure the performance of real-time DMN detection algorithms against a comparable offline method established in the literature that is not real-time compatible. This comparable offline method may be considered an emerging gold-standard in the research space for detecting DMN activation from non-invasive neurophysiology data. The real-time DMN detection algorithms tested are implemented through proprietary software produced by Resonait Medical Technologies Pty Ltd, fully outlined in[14]. The established method for inferring the DMN states has been the subject of numerous publications [7], [10], [15], [16]. It was applied to the LEMON dataset using the open source OSL-Dynamics toolbox [17].

The offline method involves an initial set of steps for data preprocessing. EEG data was pre-processed through a pass band of 1 to 45Hz and down sampled to 250 Hz. Data was then co-registered to each subject’s structural MRI scan using a multiple local sphere forward head model and then underwent source space reconstruction using an LCM-V Beamformer projected to an eight millimetre grid. This grid was then parcellated into 38 regions of interest (ROIs) [15]. Results for this work was generated using both the raw sensor and source space data. However, using source space data would not be feasible in real-time applications given the processing pipeline.

With all data projected into a common source space, the method then fits a Time delay Embedded Hidden Markov Model (TIDE-HMM) [9]. This framework [11] fits a generative model that describes the data *X*_*t*_ at each time point t as being generated from a latent state variable *Z*_*t*_∈[*1,2,…K*], where *K* is the total number of states. For consistency with the literature [9], [11], we selected a value of *K=12*.

The complete model specification includes both the probabilistic model of temporal dynamics and the observation model, where a probabilistic model determines the activation of a specific latent state based on the input data. As described in Vidaurre et al. (2018) [9], the observation model applied uses a temporal embedding with a zero-mean Gaussian observation model:

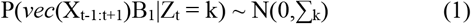

Temporal embedding was performed by the *vec* operator, where *X*_*t-l:t+l*_ is a matrix of points with dimensions [*W×P*] centred at time point *t, P* is the number of dimensions (EEG channels in this work) and *W=2l+1* is the length of the temporal embedding. The matrix *B* of dimension [*WP×Q*] reduces the dimensionality of the data for computational reasons by applying PCA with eigenvalue normalisation, with the parameter Q set to explain *90%* of the embedded data variance. For a real-time application, this model was modified to include only current and previous time points:

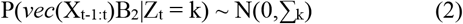

In this model *W=l+1* (where *l* was chosen to be seven samples in this work) and the [*W×P*] matrix of X_t-l:t_ points were stacked into a [*W*.*P×1*] vector through the *vec* operator. This vector was then projected to a lower dimensionality space by the linear operator *B* and modelled with a Gaussian distribution, with zero mean and a [*Q×Q*] covariance matrix determined by the active state. As a result, each RSN-state is parameterised by a unique covariance matrix, which reflects the autocovariance of each channel and the cross-covariance across channels [11]. Vidaurre et al. (2018) described how this effectively represented a parameterisation of the power spectrum and the cross-power spectrum, respectively [10], [11]. Therefore, each RSN-state corresponds to a distinct distribution of power on each channel and coherence between channels [10].

Given the large datasets and volume of data, scalable model inference was achieved using stochastic gradient variational Bayes methods that iteratively develop a model through batch training [10], [11]. [9]. To ensure convergence, full model inference was run five times, where the model with the lowest final free energy was used [11]. For RSN-state labelling, states were assigned according to ordinal distances as measured by the transition matrix and probability of direct state transitions (where high probabilities describe small distances and detailed further in Higgins et al. 2021) [11]. An active DMN state at time t would be denoted by Z_t_=1, where the DMN corresponds to RSN-state-1.

The LEMON dataset was cross-validated using five folds, where at each fold a random non-stratified training and testing set are defined (88 and 22 participants, respectively). For the purposes of evaluation, the number of participants from the training set are adjusted to train the model and measure their performance on the testing set. These models are trained and tested using the raw sensor space data. The DMN detection accuracy was calculated, along with the DMN sensitivity and specificity. Fractional occupancy (FO) was also measured to compare the correlation from the baseline and calculated RSN for each recording. Lastly, DMN sensitivity and specificity was compared for instances of DMN with varying durations.

## IV. Results

Accuracy of DMN state detection was first measured independently over time samples, at the sampling period of 0.004 seconds. At this temporal resolution when applied real-time in sensor space, the model achieved a DMN detection accuracy of 95%, with a sensitivity of 45% and specificity of 97% (see Table I).

**TABLE I.** Cross-validated DMN performance

|  | Accuracy | Sensitivity | Specificity |
| --- | --- | --- | --- |
| Sensor | 0.95±0.018 | 0.45±0.16 | 0.97±0.018 |

Comparing these results to the detection accuracies of other states in sensor space shows that the DMN state has the third highest accuracy and sensitivity of any state. This supports the conclusion that the DMN state profile, consisting of elevated alpha band power and coherence distributed over posterior cingulate and lateral-parietal brain regions concurrent with elevated delta/theta band power and coherence across frontal-temporal regions, is relatively well predisposed to real-time detection compared to other brain networks.

We then investigated how DMN sensitivity and specificity related to traits of individual DMN state visits. Fig 2 plots the sensitivity of detection vs state visit length, alongside the specificity as a function of inter-state interval length. This demonstrates a robust relationship, where the real-time detection performance is much more sensitive for longer state visits (rho=0.922, p=7.1E-32), reaching an average sensitivity of 70% for state visits over 300 milliseconds in length (see Fig 2). Conversely, the real-time detection performance displays higher specificity over inter-state intervals that are longer in duration (rho=0.885,p=6.95E-51), reaching around 90% for intervals exceeding 300 milliseconds. Taken together, these results suggest real-time performance is more robust to both intervals and state visits that are sustained over time, traits which have been attributed to stronger correspondence to cognitive processes [13].

**Fig. 1.**
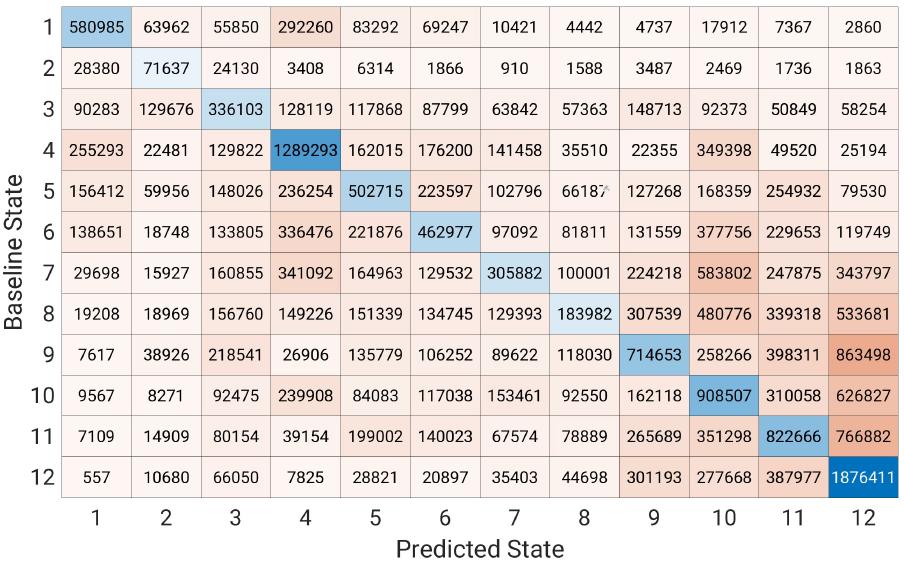
Confusion matrix of five-fold cross-validated results; state 1 is the DMN state.

**Fig. 2.**
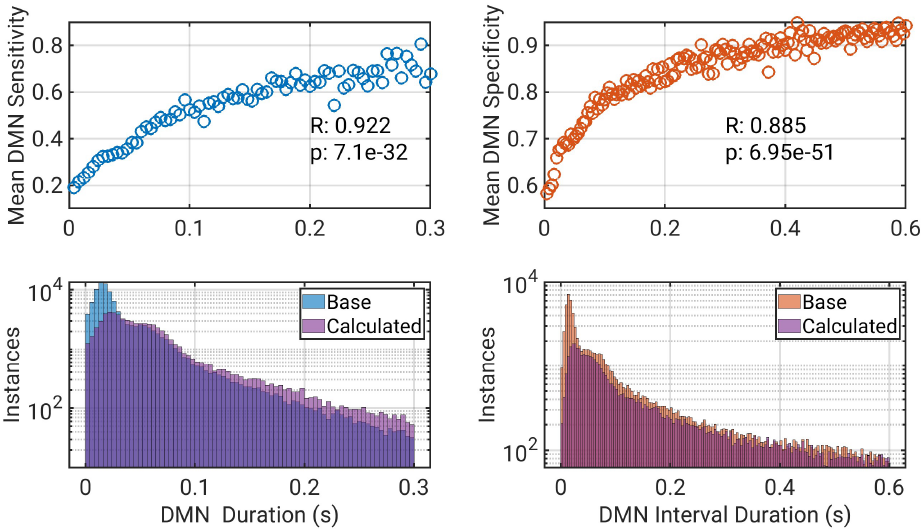
Real-time performance improves over state visits and intervals that are sustained over time. DMN sensitivity increases very robustly with state visit duration (R=0.922, p=7.1E-32). Concurrently, DMN specificity increases very robustly with inter-state interval duration (rho=0.885,p=6.95E-51).

We then investigated how well the algorithm can detect differences in each individual’s baseline state profiles, which could be important for a variety of prognostic or diagnostic applications. The fractional occupancy over all inferred states was measured and compared across subjects. This achieved a correlation of R=0.617 (p=7.11E-13 see Fig 3). The Bland-Altman plots also revealed good agreement between the baseline and calculated fractional occupancy, albeit with a small negative bias. However, this bias might be explained by the poor detection of DMN states with short durations.

**Fig. 3.**
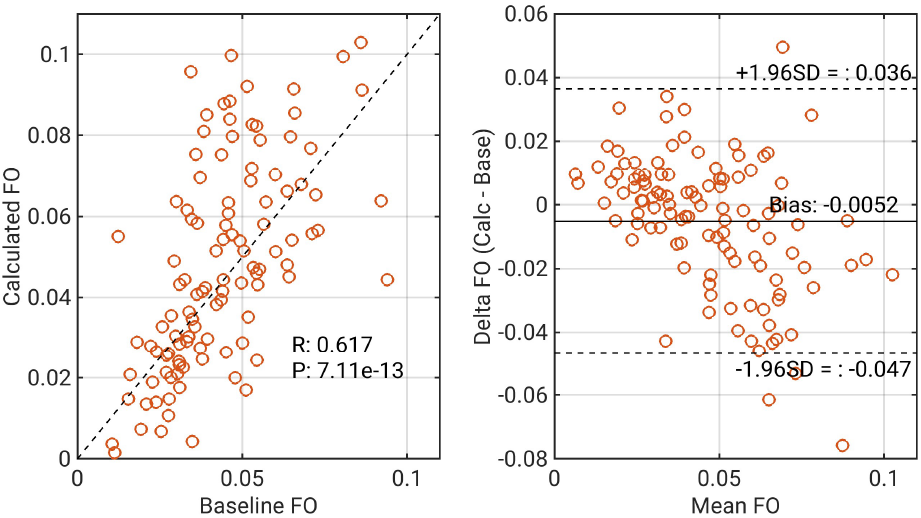
Correlation of fractional occupancy (FO) of baseline and calculated cross-validated results with corresponding Bland-Altman plots.

## V. Discussion

An ability to detect activity in the brain’s default mode network in real-time could have a range of potential clinical applications for mental health illnesses, particularly depression. The results presented above validate the technical accuracy of the real-time system tested, highlighting the potential to measure DMN activity from real-time EEG recordings. In particular, a range of metrics presented comparing accuracy of DMN detection firstly at the temporal resolution of 0.004sec, then over longer timescales, and then across different subjects supports the conclusion that DMN detection performance is reasonably robust across different timescales and comparisons.

Nonetheless, some caution is warranted interpreting these results. Firstly, our analysis assumes the output of the HMM-TIDE model on source reconstructed data is an objective ground truth. This is a modelling assumption, and in reality no underlying ground truth for the time-course of DMN activation is obtainable. Brain networks are defined by patterns of correlations over disparate anatomical regions, which do not have discretised boundaries in either time or space. The emerging gold standard is to impose such boundaries in a data-driven way, where these boundaries are inferred from a statistical analysis of a large dataset. This also requires that data is co-registered to an individual’s MRI and EEG fiducial markers, correcting for individual differences in neuroanatomy, then source reconstructed to a common brain space. Our results should be interpreted therefore as measuring concurrence with this approach, as comparison to an objective ground truth is not possible.

In a similar manner, when computing accuracy metrics we conservatively treat all states as independent. In reality brain networks are not independent but overlap in both function and physiology. The confusion matrix presented in Fig 1 for example shows the most common misclassification for DMN false negatives was into state 4, whereas the least common misclassification was into state 12. Our performance metrics conservatively weight each of these misclassifications equally, yet state 4 is the Parietal Alpha Network that is functionally linked to the DMN [11], whereas state 12 is the Dorsal Attention Network that is robustly anticorrelated with the DMN [18].

The pattern between accuracy metrics and timing patterns is also encouraging when considering the practical applications of this algorithm. Neurofeedback involves delivering a stimulus to a user when a brain state is detected, however the latency between a stimulus being triggered and it actually being transmitted to the relevant areas of the brain is on the order of 200-300 milliseconds [19]. The temporal profile of detection accuracy is quite serendipitous in this regard, being more strongly weighted to trigger selectively during longer DMN state visits, and similarly less likely to trigger a false negative during longer intervals between DMN state visits. Such a profile increases the likelihood of the stimulus reaching the relevant brain areas while the network is still engaged, despite the latency involved.

Similarly, the detection of each instance of DMN might not be as valuable as the ability to capture each individual’s FO of DMN, which may prove more clinically relevant. The Bland-Altman plot demonstrates the ability to correctly predict the FO for each participant with a high correlation (R=0.617) and a weak negative bias (see Fig 3). The demonstrated ability to detect and quantify DMN activity will form the basis for future work to detect and evaluate patients with depression and psychiatric disorders.

## VI. Conclusion

These results have demonstrated the ability to use a sub-selection of the LEMON dataset to predict the brain-state time course using the Resonait model. The clinically important DMN state was detected with a high degree of accuracy (95%) and specificity (97%), but with considerably lower sensitivity (45%). The source of missed DMN instances appears to be associated with occurrences of DMN with very short durations, which may be less clinically meaningful for therapeutic applications. The results furthermore confirm a high correlation in the FO statistics over subjects, achieving an R-value of 0.617. Taken together, these results validate the approach of real-time DMN detection in EEG data, which has potential for a range of clinical applications in mental health.

## Data Availability

All data analysed in this study are available online at: https://fcon_1000.projects.nitrc.org/indi/retro/MPI_LEMON.html

https://fcon_1000.projects.nitrc.org/indi/retro/MPI_LEMON.html

